# BRAIN RE-IRRADIATION IN LUNG CANCER – NOT AN EXERCISE IN FUTILITY

**DOI:** 10.1101/2020.04.15.20061879

**Authors:** Jai Prakash Agarwal, Shreyasee Karmakar, Anil Tibdewal, Naveen Mummudi

## Abstract

**BACKGROUND:** Whole brain radiation therapy (WBRT) is an effective palliative measure and provides durable symptom relief in lung cancer patients with multiple brain metastases (BM). Clinico-radiological progression of BM after WBRT is a common and challenging scenario; treatment is tailored, with various factors like driver mutation status, age, performance status, progression free interval and time since last irradiation influencing the treatment decision. Surgery or focal RT with stereotactic techniques may be an option for patients with oligo-metastases. However, they might not be a feasible option for patients with multiple BM. We aim to study the impact and outcome of patients with BM from lung cancer receiving re-WBRT for clinico-radiological progression.

**MATERIALS AND METHODS:** We retrospectively analyzed patients with BM from lung cancer who were registered at Tata Memorial Hospital, Mumbai, India between January 2016 to January 2019 and had undergone two courses of WBRT. Data of patients were retrieved from electronic medical records. Patients were treated using conventional or conformal technique with either tele-cobalt or Linear accelerator.

**RESULTS:** Out of 446 patients with lung cancer, diagnosed and treated with WBRT for BM, 6% patients (n=28) received re-WBRT. There were 16 men and 12 women with a median age of 53 years (range 30 to 70 years). Primary histology was adenocarcinoma in all except two patients who had small cell histology. Eighteen patients had driver mutation positive disease (11 with EGFR and 7 with ALK mutation) and a majority of patients (54%) had BM at presentation. Clinico-radiological progression was the commonest indication of re-WBRT. A majority of these patients had developed new symptoms while about 25% had recurrence of previous symptoms. Mean Karnofsky performance score (KPS) prior to re-WBRT was more than 70 in 13 patients (57%). Mean time interval between the two courses of WBRT were 16 months (range 5-37 months). Most patients received WBRT using a conventional technique (91%) and were treated in a tele-cobalt unit (83%). Re-WBRT fractionation schedule was 25 Gy/10 fractions (n=17, 61%) or 20 Gy/5 fractions (n=10, 36%). Mean biological effective dose (BED_2Gy_) for the first and second courses of WBRT were 63Gy and 56Gy respectively. The average cumulative BED_2Gy_ was 118.91Gy (range 116.25 – 120Gy). Almost all patients received short acting steroids during the course of re-WBRT. All patients except for one completed the course of treatment. At a median follow up of 2.5 years, median survival of patients after re-WBRT was 5 months. Median survival since re-WBRT was 8 months if pre first course of WBRT ds-GPA was 3.5-4 vs 1 month if it was 0-1 (p= 0.025).

**CONCLUSION:** In lung cancer patients with symptomatic progression of multiple BM and good prognostic features (driver mutation positive, good performance status and long time interval since last WBRT), re-WBRT is safe and associated with better outcomes.

## INTRODUCTION

Brain metastasis is one of the most common sites of distant metastasis in cancer patients and 20-40% of patients diagnosed with cancer are estimated to develop brain metastases(1).Almost 16-20% of non-small cell lung cancer develop brain metastasis and around 10% patients present with upfront metastasis. Incidence of brain metastasis is higher in EGFR mutated and ALK re-arranged NSCLCs;24.4% at diagnosis and 46.7% at 3 years in EGFR-mutated and 23.8% at diagnosis and 58.4% at 3 years in ALK-rearranged NSCLCs (2). Prior studies have reported intracranial relapse after whole brain irradiation in around 47-86% of patients (3).In small cell lung cancer (SCLC) patients treated with prophylactic cranial irradiation (PCI), brain recurrence is reported to be around 10-15%(4)(5).

Whole brain radiation therapy (WBRT) provides temporary palliation of symptoms, improves quality of life(6) and also survival (3)(7)(8). In the QUARTZ trial where patients were randomized between optimal supportive care with dexamethasone vs dexamethasone with WBRT, improved survival was seen in patients <60years of age, good performance status and controlled primary (9). Application of QUARTZ trial in clinical practice has shown that gender, performance status and EGFR mutation had impact on survival (10). However, almost half of the patients treated with WBRT gradually develop progressive intracranial disease(11). The median survival of lung cancer patients with brain metastases is around 4 months (12). With the advent of novel therapeutics like targeted therapy for driver mutation positive lung cancer patients, survival of patients with brain metastasis has improved(13). As such, there are more patients warranting treatment for progressive brain metastases. Re-irradiation (re-RT) in such patients can be offered using WBRT, partial brain radiotherapy (RT) or high dose conformal stereotactic radiotherapy (SRT). But considering tolerance dose of brain and weighing the risk-benefit ratio of re-RT, only 3-10% of patients are offered re-RT(11).

Though there have been several studies(3)(14)determining the outcome of re-RT in patients with brain metastases, those studies included patients with metastases from various primary sites-breast cancer being the most common cause. The aim of our study is to determine outcomes of lung cancer patients receiving re-irradiation with WBRT (re-WBRT) for brain metastases and to find correlation between outcome and various clinical features.

## MATERIALS AND METHODS

### Source Of Data

We selected patients from a prospectively maintained database which comprised of patients with brain metastases from lung cancer who received re-WBRT at our institution, between January 2016 and January 2019. Data regarding patient, disease and treatment-related variables were collected from electronic medical records and treatment planning systems. The study was approved by the Institutional review board and consent was waived.

### Patients

Patients who had histopathological proof of lung cancer, age > 18 years and had received re-irradiation for brain metastases were included in the study. Patients treated outside our institution were also included in the study provided that complete treatment details were retrievable. Karnofsky performance score (KPS), disease specific graded prognostic assessment (DS-GPA) and recursive partition analysis (RPA) were noted before initial RT and re-RT. Epidermal growth factor receptor (EGFR) mutation and anaplastic lymphoma kinase (ALK) mutation status were also documented.

### Interventions

All patients had received two courses of radiotherapy to whole brain. These included patients receiving either prophylactic cranial irradiation (PCI) or whole brain RT with palliative intent. The decision regarding first course of RT and re-WBRT were decided based on either clinical symptoms or imaging findings. Neuro-imaging was done using either computed tomography scan (CT scan) or magnetic resonance imaging (MRI). Treatment was delivered on a linear accelerator or on a tele-cobalt therapy unit to whole brain with appropriatemargins to account for set-up errors. A total dose of 20-30 Gy in 5-10 fractions at 3-4Gy per fraction over 1-2 weeks was delivered using either conventional or 3-dimensional conformal techniques (3DCRT) based on clinical decision.

After completion of radiotherapy, patients were followed up at regular intervals with clinical examination and repeat imaging when indicated.

### Outcomes

Overall survival (OS) of patients and survival of patients since re-WBRT were calculated and their association with driver mutation status was analyzed.

### Statistical Analysis

Overall survival of patients was calculated from date of registration to date of death or date of last follow-up whichever was later. Survival of patients since re-WBRT was calculated from last date of re-WBRT to date of last follow-up or date of death whichever was later. All analysis were done using SPSS version 21. Patient characteristics were described with median and extreme values for continuous variables and with frequencies and percentages for categorical variables. Overall survival and survival since re-WBRT was plotted using Kaplan-Meier survival curves. Log-rank test was used to determine association between driver mutation status and survival.

## RESULTS

We retrieved data of 446 consecutive patients of lung cancer with brain metastases who had received whole brain radiotherapy. Of these, 28 (6.3%) patients had received re-WBRT. The patient age ranged from 30-70 years, median age being 53 years. More than half of the patients were male (n=16, 57.1%). Most of the patients did not have any habits and only 14.3% of the patients were smokers. Table 1 lists the demographic features of the patient population.

**Table 1:** Patient demographics.

| Parameter |  | Number (%) |
| --- | --- | --- |
| Age | 53 years ( Range- 30- 70 years) |  |
| Sex | Male | 16 (57.1%) |
|  | Female | 12 (42.9%) |
| Habits | Smoking | 4 (14.3%) |
|  | Tobacco chewing | 3 (10.7%) |
|  | None | 21 (75%) |
| Histology | Adenocarcinoma | 25 (89.3%) |
|  | Small cell | 2 (7.1%) |
|  | Small cell + Adeno | 1 (3.6%) |
| EGFR status | Mutated | 11 (39.3%) |
|  | Non-mutated | 17 (60.7%) |
| ALK status | Positive | 7 (25%) |
|  | Negative | 21 (75%) |
| EGFR- Epidermal growth factor receptor; ALK- anaplastic lymphoma kinase |  |  |

### First course of WBRT

At baseline, the median KPS of patients were 80 (range 40-90). Patients were subdivided into groups with KPS < 70 and KPS >/= 70. Most of the patients were of class II RPA. Half of the patients had ds-GPA of 1.5-2 (n= 14, 50%), as depicted in table 2

**Table 2:** Performance status of patients.

| Parameter |  | At first course of WBRT | At re-WBRT |
| --- | --- | --- | --- |
| KPS | <70 | 7 (25%) | 12 (42.9%) |
|  | =/ > 70 | 21 (75%) | 16 (57.1%) |
| RPA | Class i | 4 (14.3%) | 1 (3.6%) |
|  | Class ii | 14 (50%) | 12 (42.9%) |
|  | Class iii | 10 (35.7%) | 15 (53.6%) |
| DS-GPA SCORE | 0-1 | 4 (14.3%) | 6 (21.4%) |
|  | 1.5-2 | 14 (50%) | 21 (75%) |
|  | 2.5-3 | 7 (25%) | 1 (3.5%) |
|  | 3.5-4 | 3 (10.7%) | 0 |
| KPS- Karnofsky performance score; RPA- Recursive partition analysis; ds-GPA- disease specific graded prognostic assessment |  |  |  |

Around one-third patients had metastasis in brain only while 64.3% (n=18) had metastasis in brain as well as extra-cranial site. Among the patients, 53.6% (n=15) had presented upfront with brain metastasis while the remaining patients (n=13, 46.4%) developed brain metastasis during the course of their treatment. Headache was the most common symptom at initial presentation (n=12, 42.9%), followed by seizure (n=6, 21.4%) and weakness (n=6, 21.4%). Only 32.1% (n=9) patients had neuro-deficit at presentation. Three patients with small cell lung cancer (SCLC) received prophylactic cranial irradiation. Seven patients (25%) received WBRT due to radiological indication.

Twenty-five patients (89.3%) patients had adenocarcinoma, 2 (7.1%) had SCLC and 1 (3.6%) had both adeno and small cell component. EGFR mutation was present in 39.3% (n=11) patients and only 25% (n=7) patients were ALK positive.

All patients had undergone imaging prior to first course of WBRT. MRI was done in 22 patients (78.6%) and 3 patients (10.7%) underwent contrast-enhanced CT (CECT) scan of brain. In 3 patients, brain metastasis was detected in PET-CECT scan done as a part of metastatic work-up. Majority of the patients (n=21, 75%) had multiple brain metastases, 17.9% patients (n=5) had single metastasis and 2 had none (prophylactic cranial irradiation).

Almost all patients received radiation to whole brain using conventional techniques except 1 who had received WBRT using 3DCRT. Twenty-one patients (71.6%) were treated with cobalt 60 gamma rays and 6 were treated with 6 MV photons in a linear accelerator. Dose delivered was 20-30Gy delivered at 3-4Gy per fraction over 1-2 weeks. The Mean biological effective dose (BED_2Gy_) was 62.94Gy.

### Re-irradiation

The median KPS of patients was 70 (range 30-80). Fifteen patients were of RPA class III and 75% patients had ds-GPA 1.5-2 as depicted in table 2. Majority of the patients (n=18, 64.3%) had developed new symptoms post initial WBRT necessitating re-WBRT. Only 7 patients (25%) had recurrence of similar symptoms as that prior to first course of WBRT and 2 had both new as well as recurrence of symptoms. Twenty-two patients (78.6%) received re-WBRT due to clinical as well as radiological progression. Contrary to initial presentation, the most common presenting symptom was weakness (n=12, 42.9%). Seven patients (25%) had headache and 5 patients (17.9%) had presented with seizure. Radiological evidence of progression was seen in 89.3% patients.

Twenty patients (71.4%) had undergone MRI prior to starting re-WBRT, 5 patients (17.9%) had CECT brain while 3 patients did not have any imaging and was started on re-WBRTbased on clinical judgement. Three patients had single brain metastasis which had progressed from before, 1 patient had developed leptomeningeal disease and 21 patients had multiple brain metastasis.

### Re-WBRT details

Median time interval between re-WBRT and first course of WBRT was 16 months (range 5-37 months). Most of the patients (n=23, 82.1%) were treated with cobalt 60 gamma rays and 5 (17.9%) patients were treated with 6MV photons in linear accelerator. Two of these five patients were treated with 3DCRT while rest of the patients were treated with conventional technique. Re-WBRT fractionation schedule was 25 Gy in 10 fractions (n=17, 61%) or 20 Gy in 5 fractions (n=10, 36%)’ one patient received 12Gy in 2 fractions once weekly. Mean biological effective dose (BED_2Gy_) for re-WBRT was 55.97Gy (range 56.25 – 60Gy). The average cumulative BED_2Gy_ was 118.91 Gy (range 116.25 – 120Gy).

### Compliance to radiation, toxicities and steroid usage

There was good compliance to treatment and all patients completed WBRT. At the time of re-irradiation, radiation was stopped early in one patient after completion of 16Gy delivered in 4 fractions at 4Gy per fraction due to poor tolerance and poor general condition. All other patients completed re-WBRT with good compliance.

Patients had minimal toxicities both post first course of WBRT and post re-WBRT. Post first course of WBRT, 2 patients had developed grade I skin reaction, 1 patient had decreased memory. Even after re-WBRT, only 2 patients (7.1%) had developed grade I skin toxicity and rest did not develop any toxicity. Almost all patients had received steroids in tapering dose ranging from 12-24mg daily in 2 to 3 divided doses during both courses of radiation.

### Clinical and radiological benefit

Post first course of WBRT, partial or complete relief of symptoms was seen in 7 patients. After completion of re-WBRT, 8 patients had complete or partial symptomatic relief, one patient had increased drowsiness while one had increased neurological deficit.

Post re-WBRT, 7 patients had undergone response assessment MRI and 1 had undergone CT scan. Twenty patients had not undergone any imaging. Of the patients who had undergone imaging, 3 patients had stable disease, 4 had progressive disease and 1 had decrease in size and number of metastases. Radio-necrosis was not seen in any of the patients.

### Survival outcomes

At a median follow-up of 2.5yrs, the median survival since re-WBRT was 5 months. There was no statistically significant survival since re-WBRT difference based on gender, age (<60yrs vs >/= 60yrs) and presence of extra-cranial disease at presentation. There was no statistically significant difference in median survival since re-WBRT between EGFR mutated, ALK positive and EGFR and ALK negative cohort (7 months vs 5 months vs 2 months respectively, p= 0.43) (Fig 2).

**Fig 1.**
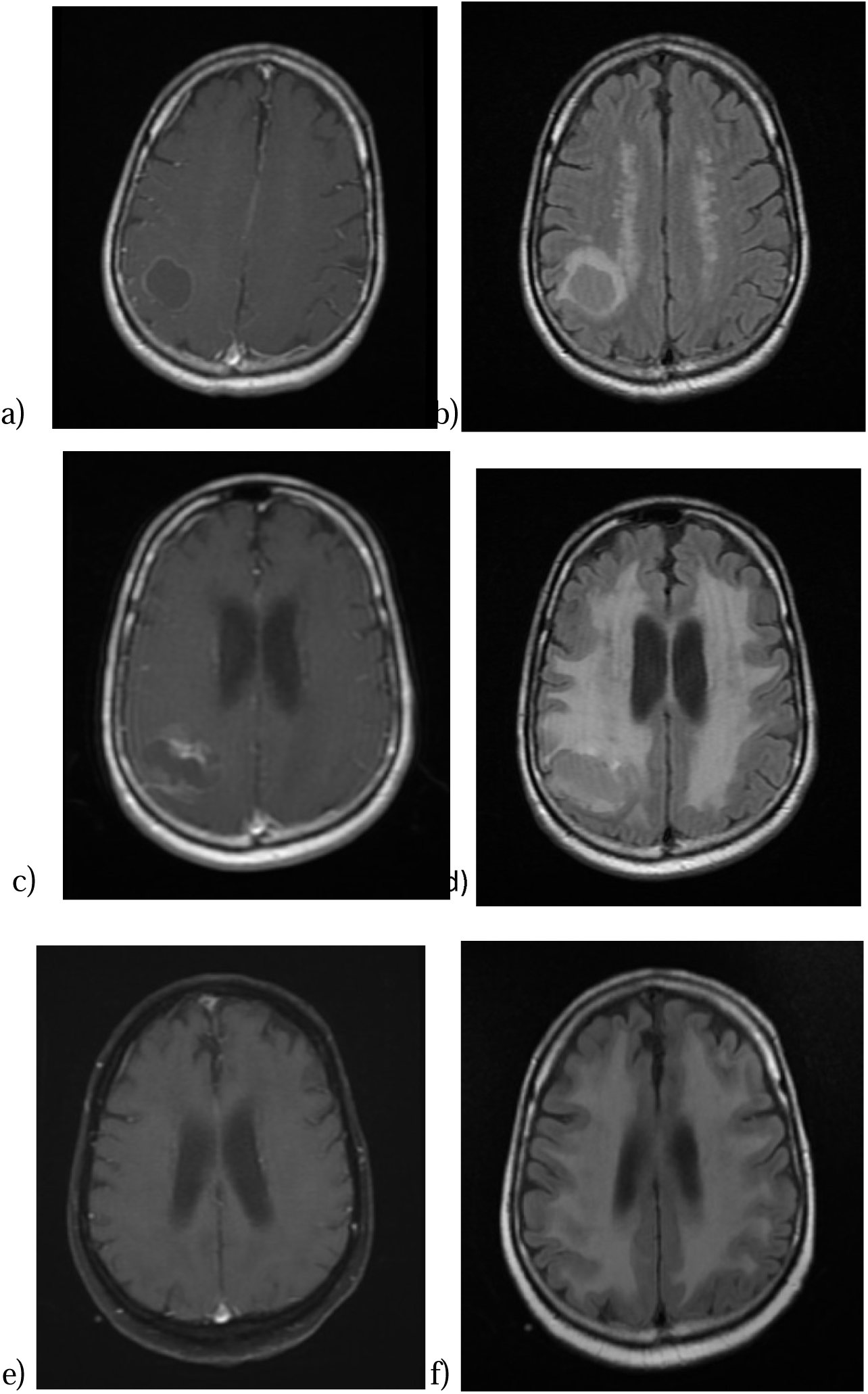
(a, b)- T1contrast and T2 flair MRI at presentation; (c, d)-T1 contrast and T2 flair MRI images post WBRT at progression; (e, f) - T1 contrast and T2 flair MRI images post Re-WBRT showing controlled disease with no radio-necrosis.

**Fig 2:**
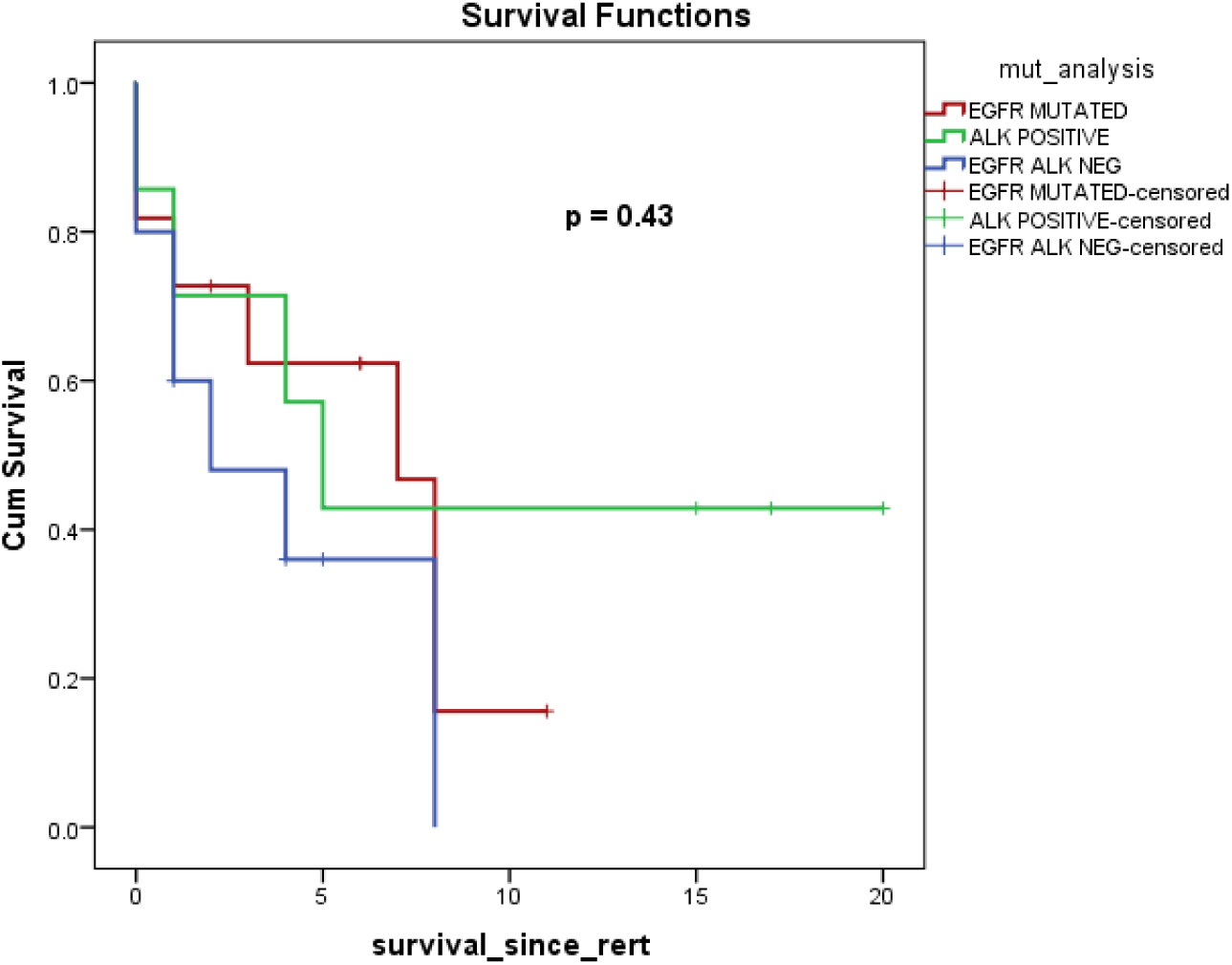
Survival since re-WBRT in EGFR mutated vs ALK positive vs EGFR ALK negative.

It was seen that overall survival since re-WBRT was better if RPA and ds-GPA prior to first course of WBRT were more. Median survival since re-WBRT was 8 months if pre first course of WBRT ds-GPA was 3.5-4 vs 1 month if it was 0-1 (p= 0.025). In patients with pre first course of WBRT ds-GPA >2, median survival since re-WBRT was 8 months while in those with ds-GPA < 2, it was 2 months only (p=0.04) (Fig 3a). Survival was better in patients with RPA class I prior to first course of WBRT (p=0.002). There is a trend of better survival in patients of class I RPA at re-WBRT. However, ds-GPA at re-WBRT did not affect survival since re-WBRT.

**Fig 3:**
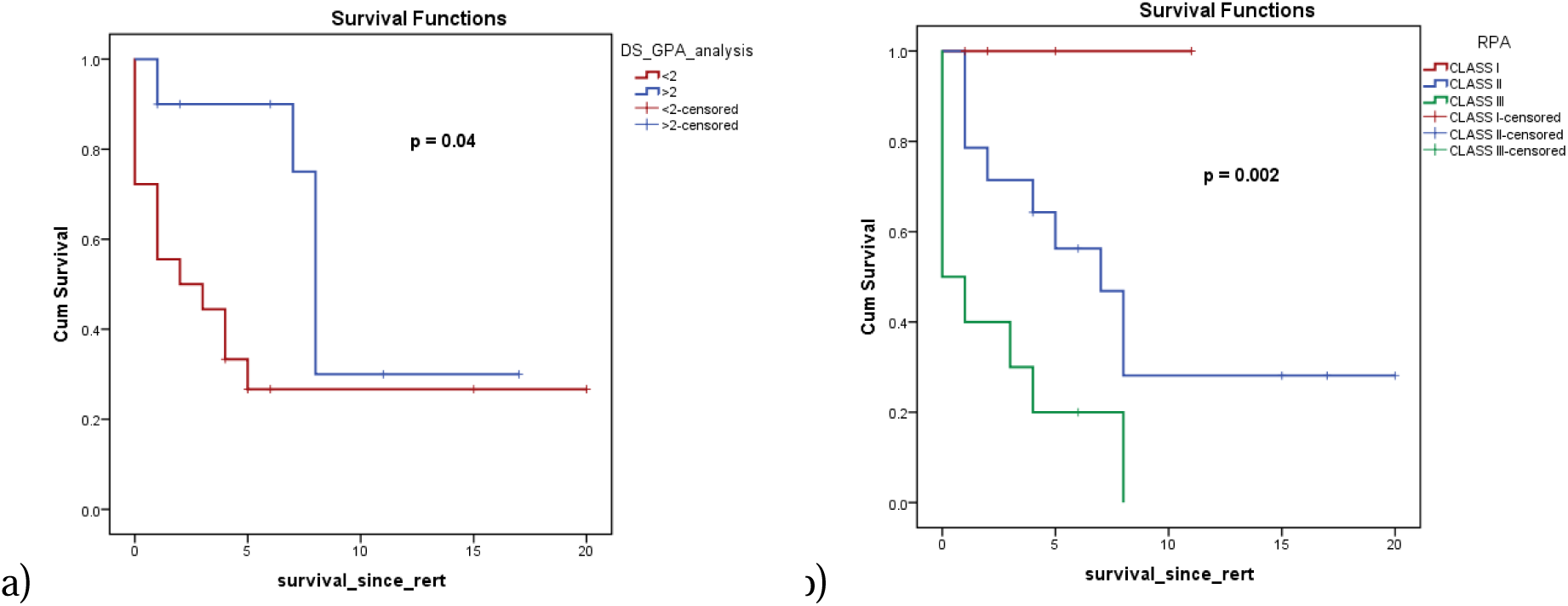
(a) Survival since re-WBRT based on pre first course of WBRT ds-GPA< 2 vs ds-GPA >2. (b) Survival since re-WBRT based on pre first course of WBRT RPA

**Fig 4:**
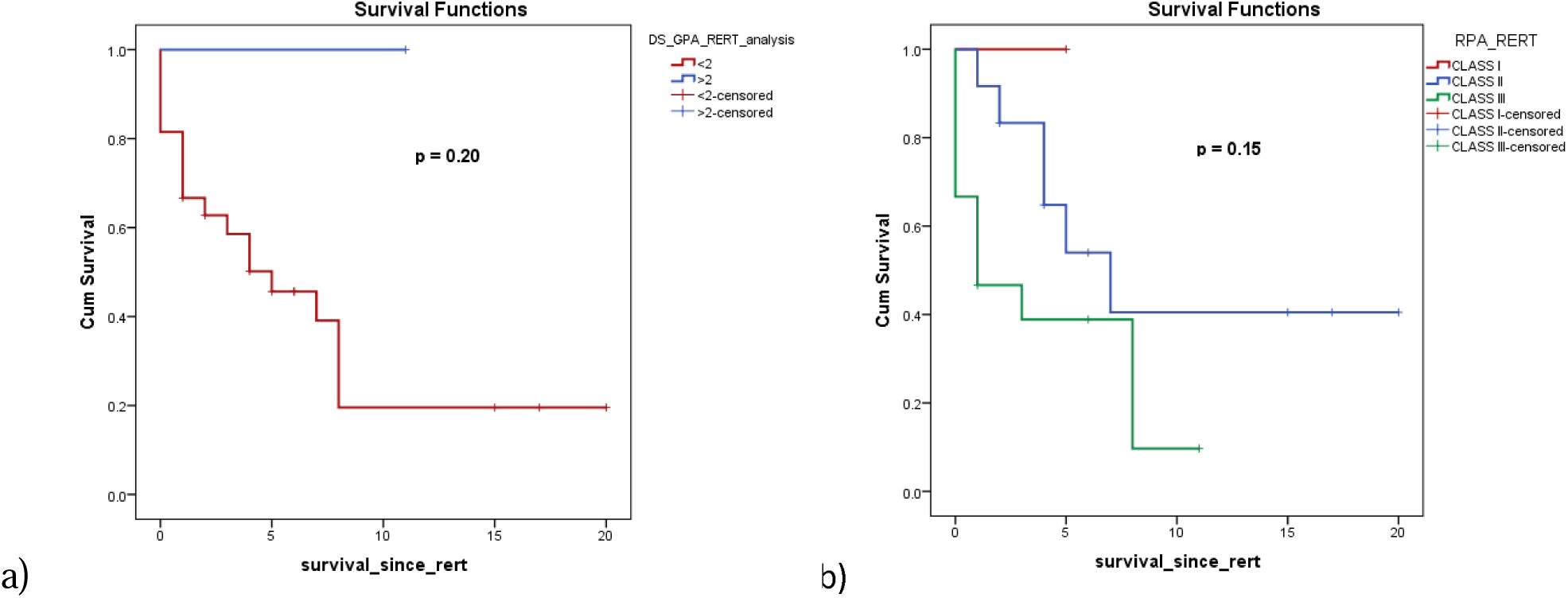
(a) Survival since re-WBRT based on re-WBRT ds-GPA. (b) Survival since re-WBRT based on re-WBRT RPA.

There was no statistically significant difference in survival since re-WBRT based on KPS pre-first course of WBRT (8.8 months for KPS >/=70, 3.2 months for KPS < 70, p=0.1) and KPS at re-WBRT (9.8 months for KPS >/=70, 4.2 months for KPS < 70, p=0.08).

## DISCUSSION

Re-irradiation for symptomatic progression of brain metastases is often a matter of debate and there is no consensus as to which patients should ideally receive re-WBRT. The American College of Radiology recommends that management options should be based on patient characteristics and preferences, previous treatments and potential risks and toxicities of treatment(15). Though WBRT palliates symptoms and improves quality of life of patients, re-WBRT is often considered challenging owing to the risk of radio-necrosis and neurocognitive decline. In our study, we had irradiated 28 patients of lung cancer with brain metastases for either clinical or radiological progression post first course of WBRT and we found re-WBRT for clinically progressive brain metastasis in lung cancer patients to be a safe and feasible option. At a median follow-up of 2.5yrs, the median survival after re-WBRT in our study was 5 months. *Aktan et al*(16) reported median survival of 5.3 months post re-irradiation and *Akiba et al*(14) found median survival post re-WBRT as 4 months. The median survival after re-WBRT reported by various other studies also range from 2.5-5 months(7)(17)(18) and this corroborated with findings of our study. The benefit of re-WBRT was seen more in patients with good RPA and ds-GPA prior to first course of WBRT. However, RPA and ds-GPA at re-WBRT seemed to have lesser impact on survival since re-WBRT, though a trend towards better survival was observed in patients of class I RPA at re-WBRT. Post re-WBRT patients had minimal toxicities and all had completed treatment except one in view of poor general condition.

Several retrospective studies have evaluated the effect of re-WBRT with various dose and fractionation schedules at. *Shehata et al* in their study had treated majority of the patients to a dose of 10Gy single fraction. *Aktan et al*(16) had used a median dose of 30Gy in 15# for the initial WBRT. Same dose schedule was observed by several other retrospective studies. The median dose delivered in our study at first course of WBRT was 20Gy in 5# at 4Gy per fraction delivered over 1week. *Aktan et al* used re-WBRT dose as 25Gy in 10# in majority of patients which was similar to that used in our study. They achieved a cumulative BED_2Gy_ of almost 130Gy. *Akiba et al*(14) in their retrospective study of 31 patients had achieved a cumulative BED_2Gy_ of 228Gy. In our study was, cumulative mean BED_2Gy_ of re-WBRT was 116.25Gy (see table 3).

**Table 3:** Comparison of various studies and outcomes.

| **AUTHOR** | **NO. OF PATIENTS** | **PRIMARY** | **WBRT DOSE** | **RE-WBRT DOSE** | **INTERVAL** | **MEDIAN SURVIVAL SINCE RE-WBRT** | **TOXICITIES** |
| --- | --- | --- | --- | --- | --- | --- | --- |
| Shehata et al, 1974 (23) | 35 |  | 1X10Gy<br>10X3Gy | 1X10Gy | - | - | 17% acute toxicity |
| Kurup et al, 1980 (24) | 56 | Lung, breast | 3X6Gy | 10X2Gy | 6.3months<br>(mean) | 3.5months | 18% acute toxicity |
| Cooper et al, 1990 (18) | 52 |  | 10X3Gy | 10X2.5Gy | >4 months | 4 months | - |
| Wong et al, 1996 | 86 | Breast, lung, colon, melanoma, others | 10X3Gy | 10X2Gy | 7.6 months<br>(median) | 4 months | 5 patient with radiologic abnormality |
| Son et al, 2012 (17) | 17 | Lung, breast, colorectal | 14X2.5Gy | 12X1.8Gy | 15 month<br>(median) | 5.2 months | 71% with acute side effect |
| Scharp et al, 2014 (21) | 134 | Lung, breast, melanoma, others | 15X2Gy | 10X2 Gy | 13.4 months<br>(median) | 2.8 months | 67% with acute side effects, 10 did not complete treatment |
| Aktan et al, 2015 (16) | 34 | Lung, breast, others | 10X3Gy | 10X2.5Gy | 12.8 months<br>(median) | 5.3months | - |
| Present study | 28 | Lung | 5X4Gy<br>10X3Gy | 10X2.5Gy | 16 months<br>(median) | 5.1months | 7.1% acute skin reaction |

Absence of extra-cranial disease, young age and better performance status (KPS>70) has been associated with better outcomes post re-WBRT in our study and this was similar with several other studies.*Cooper et al*(18)found that patients with younger age and no extra-cranial metastases had favorable outcomes. *Wong et al*(19)saw that 12 month and 18 month survival rates of patients were significantly better in patients with no extra-cranial disease (14% vs 4% and 11% vs 0%, p=0.025). *Sadikov et al*(20) and *Aktan et al*(16) also reported no difference in overall survival in patients with respect to presence or absence of extra-cranial disease. *Scharp et al*(21) observed that patients with KPS >/= 80 had a median survival of 4.9months vs 3.1 months in patients with KPS < 80.*Akiba et al*(14)found that KPS >/=70 at the time of re-irradiation had significant better survival. *Aktan et al* also reported that patients with KPS > 70 had better survival compared to patients with KPS < 70. However, they had not reported the impact of KPS on survival since re-WBRT. We found that there was no statistically significant difference in survival since re-WBRT between KPS > 70 and KPS < 70 at either initial presentation or at re-WBRT. We only found a single study evaluating the relation of RPA with post re-WBRT survival(22). They reported that median survival was better in patients with post re-WBRT RPA class I and this was in concordance with findings of our study.

We, however, did not find any study analyzing the effects of ds-GPA and driver mutation status on survival since re-WBRT. We found that median survival since re-WBRT was significantly better if pre-first course of WBRT ds-GPA and RPA were higher. There was a trend of improved survival since re-WBRT if RPA at re-WBRT was higher. However, we did not find any statistically significant survival difference based on EGFR and ALK mutation status. This may be due to small number of patients included in the study.

The chances of toxicities associated with a second course of WBRT often precludes the clinician from offering re-irradiation in many patients. *Scharp et al*(21) had reported that almost 10% patients had to discontinue their treatment owing to the toxicities. *Wong et al*(19)did not report any severe toxicities. However, 5 patients had radiation related changes in imaging. Brain atrophy post re-WBRT as detected in MRI has been reported in around 74% of patients and cognitive disturbance or encephalopathy of grade 2 or higher was seen in around 30% patients in another study (14).*Son et al*(17)reported fatigue (35.3%), headache (23.5%), nausea/vomiting(23.5%), ataxia (5.9%), skin irritation (5.9%), and dizziness (5.9%).In our study we did not find any severe toxicities post re-WBRT. Maximum toxicity seen in our study was RTOG grade I skin reaction. The reason for lower acute toxicity seen in our patients might be due to better patient selection for offering re-WBRT and preserved general condition of patients. Radio-necrosis was seen in none of the patients who had undergone subsequent imaging after re-WBRT.

The strength of our study is that we have reported the use of re-WBRT in patients with lung cancer exclusively in contrast to majority of the previous studies that had taken into consideration all primary sites. Given the significantly better survival with the use of targeted therapy in patients with driver mutation positive NSCLC, it is important to assess the role of re-WBRT in this clinical scenario. To our knowledge, ours is the only study reporting the outcomes of re-WBRT with regards to EGFR mutation and ALK positivity. We also acknowledge the weakness of our study, including the retrospective nature of our study, small sample size, absence of quantitative or qualitative measures of cognitive function and the non-availability of post re-WBRT imaging in all patients.

In conclusion, we found that re-WBRT is a safe and feasible option for lung cancer patients with symptomatic or radiologic progression of brain metastases. Patients with higher ds-GPA and RPA at baseline have better survival with re-WBRT. The role of re-WBRT in EGFR mutation and ALK positivity needs to be further validated in larger cohort of patients.

## Data Availability

Data is available

## REFERENCES

1. Soffietti R, Ruda R, Trevisan E. Brain metastases: current management and new developments. Curr Opin Oncol. 2008 Nov;20(6):676–84.

2. Rangachari D, Yamaguchi N, VanderLaan PA, Folch E, Mahadevan A, Floyd SR, et al. Brain metastases in patients with EGFR-mutated or ALK-rearranged non-small-cell lung cancers. Lung Cancer [Internet]. 2015;88(1):108–11. Available from: http://dx.doi.org/10.1016/j.lungcan.2015.01.020

3. Borgelt B, Gelber R, Kramer S, Brady LW, Chang CH, Davis LW, et al. The palliation of brain metastases: Final results of the first two studies by the radiation therapy oncology group. Int J Radiat Oncol Biol Phys. 1980;6(1):1–9.

4. Bernhardt D, Adeberg S, Bozorgmehr F, Opfermann N, Hoerner-Rieber J, König L, et al. Outcome and prognostic factors in patients with brain metastases from small-cell lung cancer treated with whole brain radiotherapy. J Neurooncol. 2017;134(1):205–12.

5. Sun A, Hu C, Wong SJ, Gore E, Videtic G, Dutta S, et al. Prophylactic Cranial Irradiation vs Observation in Patients with Locally Advanced Non-Small Cell Lung Cancer: A Long-term Update of the NRG Oncology/RTOG 0214 Phase 3 Randomized Clinical Trial. JAMA Oncol. 2019;5(6):847–55.

6. Horton J, Baxter DH, Olson KB. The management of metastases to the brain by irradiation and corticosteroids. Am J Roentgenol Radium Ther Nucl Med. 1971 Feb;111(2):334–6.

7. Ozgen Z, Atasoy BM, Kefeli AU, Seker A, Dane F, Abacioglu U. The benefit of whole brain reirradiation in patients with multiple brain metastases. Radiat Oncol [Internet]. 2013;8(1):1. Available from: Radiation Oncology

8. Suzuki R, Wei X, Allen PK, Welsh JW, Cox JD, Komaki R, et al. Outcomes of re-irradiation for brain recurrence after prophylactic or therapeutic whole-brain irradiation for small cell lung Cancer: A retrospective analysis. Radiat Oncol. 2018;13(1):1–10.

9. Mulvenna P, Nankivell M, Barton R, Faivre-Finn C, Wilson P, McColl E, et al. Dexamethasone and supportive care with or without whole brain radiotherapy in treating patients with non-small cell lung cancer with brain metastases unsuitable for resection or stereotactic radiotherapy (QUARTZ): results from a phase 3, non-inferiority,. Lancet [Internet]. 2016;388(10055):2004–14. Available from: http://dx.doi.org/10.1016/S0140-6736(16)30825-X

10. Agarwal JP, Chakraborty S, Laskar SG, Mummudi N, Patil VM, Upasani M, et al. Applying the QUARTZ Trial Results in Clinical Practice: Development of a Prognostic Model Predicting Poor Outcomes for Non-small Cell Lung Cancers with Brain Metastases. Clin Oncol. 2018;30(6):382–90.

11. Bahl A, Kumar M, Sharma DN, Jothy Basu KS, Jaura MS, Rath GK, et al. Reirradiation for progressive brain metastases. J Cancer Res Ther. 2009;5(3):161–4.

12. Lagerwaard FJ, Levendag PC, Nowak PJCM, Eijkenboom WMH, Hanssens PEJ, Schmitz PIM. Identification of prognostic factors in patients with brain metastases: A review of 1292 patients. Int J Radiat Oncol Biol Phys. 1999;43(4):795–803.

13. Sperduto PW, Yang TJ, Beal K, Pan H, Brown PD, Bangdiwala A, et al. Estimating survival in patients with lung cancer and brain metastases an update of the graded prognostic assessment for lung cancer using molecular markers (Lung-molGPA). JAMA Oncol. 2017;3(6):827–31.

14. Akiba T, Kunieda E, Kogawa A, Komatsu T, Tamai Y, Ohizumi Y. Re-irradiation for Metastatic Brain Tumors with Whole-brain Radiotherapy. 2012;42(February):264–9.

15. Panel E, Brain O, Jared M, Elson A, Buatti JM, Chang EL, et al. American College of Radiology FOLLOW-UP AND RETREATMENT OF BRAIN METASTASES Summary of Literature Review. 2014;1–11.

16. Aktan M, Koc M, Kanyilmaz G, Tezcan Y. Outcomes of reirradiation in the treatment of patients with multiple brain metastases of solid tumorsl: a retrospective analysis. 2015;3(21):1–6.

17. Son CH, Jimenez R, Niemierko A, Loeffler JS, Oh KS, Shih HA. Outcomes after whole brain reirradiation in patients with brain metastases. Int J Radiat Oncol Biol Phys. 2012;82(2):167–72.

18. Cooper JS, Steinfeld AD, Lerch IA. Cerebral metastases: value of reirradiation in selected patients. Radiology. 1990 Mar;174(3 Pt 1):883–5.

19. Wong WW, Schild SE, Sawyer TE, Shaw EG. Analysis of outcome in patients reirradiated for brain metastases. Int J Radiat Oncol • Biol • Phys [Internet]. 1996 Feb 1;34(3):585–90. Available from: https://doi.org/10.1016/0360-3016(95)02156-6

20. Sadikov E, Bezjak A, Yi Q-L, Wells W, Dawson L, Millar B-A, et al. Value of whole brain re-irradiation for brain metastases--single centre experience. Clin Oncol (R Coll Radiol). 2007 Sep;19(7):532–8.

21. Scharp M, Hauswald H, Bischof M, Debus J, Combs SE. Re-irradiation in the treatment of patients with cerebral metastases of solid tumors: Retrospective analysis. Radiat Oncol. 2014;9(1):1–8.

22. Logie N, Jimenez RB, Pulenzas N, Linden K, Mrt T, Ciafone D, et al. Estimating prognosis at the time of repeat whole brain radiation therapy for multiple brain metastasesl: The reirradiation score. Advancesradonc [Internet]. 2017;2(3):381–90. Available from: http://dx.doi.org/10.1016/j.adro.2017.05.010

23. Shehata WM, Hendrickson FR, Hindo WA. Rapid fractionation technique and re-treatment of cerebral metastases by irradiation. Cancer. 1974 Aug;34(2):257–61.

24. Kurup P, Reddy S, Hendrickson FR. Results of re-irradiation for cerebral metastases. Cancer. 1980 Dec;46(12):2587–9.

